# Comorbidities, Behaviors, and Socioeconomic Factors and Mortality from Diseases of the Heart in New Jersey

**DOI:** 10.1101/2023.01.26.23284982

**Authors:** Matthew Guariglia, Stephen Poos, Ahmed Gawash, David F. Lo

## Abstract

Mortality from diseases of the heart claimed the lives of 186,074 New Jerseyans from 2010 to 2019. This study aims to establish correlations between each of health-related risk factors and death from heart disease in each of the six New Jersey counties. Each of the counties ranked by age-adjusted mortality per 100,000 from diseases of the heart. The six counties were divided by the least (Hunterdon, Somerset, Bergen) and greatest (Cape May, Salem, and Cumberland) mortality rates from heart disease. Additionally, this data was broken down into three main categories that include comorbidities, socioeconomic status, and behavior patterns. Each main category is further broken down into subcategories that provide further evidence of how mortality from heart disease impacts the garden state. The main outcome of death in adults over 25 from heart disease from 2010 to 2019 was correlated to 28 health factors including comorbidities, behaviors, and socioeconomic factors. An R squared value was calculated to evaluate the strength of the correlation between each factor and the outcome of mortality from disease of the heart.

## Background

Heart disease is the leading cause of mortality in the United States^1^. The CDC defines heart disease to include coronary artery disease (CAD), heart attacks, arrythmias, and heart failure. Although CAD is the most common type of heart disease, most patients first encounter is through a myocardial infarction^1^. Myocardial infarctions often result from ischemia within the cardiac muscle from spontaneous complications of atherosclerosis resulting in coronary thrombosis. This pathogenesis is attributable to a wide range of factors including endothelial dysfunction, coronary spasm, coronary embolism, tachy-/brady-arrhythmias and hypo- and hypertension^2^.

Many comorbidities such as hypertension, obesity, and diabetes mellitus contribute to the exacerbation of heart disease (CDC). It has been established that achieving hypertensive control in the black community involves addressing comorbid diseases, such as chronic kidney disease/CKD, diabetes mellitus/DM, and obesity^3^.

Comorbidities alone is a single category that contributes to mortality of heart disease, but other contributing categories exist that include socioeconomic status and behavior patterns. These three categories are ultimately interconnected and interact with one another to form a multi-factorial health crisis that continues to affect counties throughout New Jersey. More health professionals consider lifestyle as one of the most important factors affecting health in which 70% of illnesses such as cardiovascular diseases, respiratory disorders, motor-system and muscular problems and the like are directly or indirectly correlated with lifestyle^4^. However, this lifestyle or way of living is not ultimately in one’s control due to socioeconomic status. Social factors related to socioeconomic status, such as education level, occupation, or income, influence diet quality. Low socioeconomic status populations have less access to healthy foods and more access to nutrient-dense foods that are highly processed with fats, starches and sugar^5^. These nutrient-poor foods have a confirmed role in the development of non-communicable diseases such as diabetes and CAD responsible for 70% of all deaths worldwide^6^. Continuing eating these nutrient poor foods has revealed an association between poverty and obesity that may be mediated, in part, by the low cost of energy-dense foods and may be reinforced by the high palatability of sugar and fat. This economic framework provides an explanation for the observed links between socioeconomic variables and obesity^7^. Obesity prevalence is up to 33% higher than the norm in children in low-education, low-income, and higher unemployment households^8^. Consequently, cardiovascular mortality has been observed to be significantly higher among middle and low socioeconomic status groups compared to the high socioeconomic status group^9^. The consequences of these food consumption are based on the constraints of these socioeconomic status variables that ultimately affect disenfranchised populations such as the black community. These behavior patterns of eating nutrient poor foods do not only affect adults but children who grow up in these NJ county communities. If families are struggling financially, it is cheaper and logistically easier to purchase unhealthy foods. Thus, these behavior patterns become integrated into children and ultimately put them at a higher risk of mortality from heart disease starting in early adulthood.

Other behaviors playing into disease progression include physical activity. For instance, moderate evidence indicates that High Intensity Interval Training (HIIT) can improve insulin sensitivity, blood pressure, and body composition in adults with group mean ages ranging from 20 to 77 years old^10^. It must be understood that cardiovascular recovery from acute stress, but not reactivity to stress, may be a key pathway between low SES and risk for cardiovascular diseases^11^. No matter the intensity of the workout, evidence shows walking groups have wide-ranging health benefits. Meta-analysis showed statistically significant reductions in mean difference for blood pressure, resting heart rate, body fat, body mass index, total cholesterol, and VO2 max^12^. It has been established that achieving hypertensive control in the black community involves addressing higher levels of physical inactivity^3^. Inability to do so causes behavior habits that lead to health comorbidities which build through years of living a lifestyle. The intersection of socioeconomic status, medical history, and lifestyle plays a large role in mortality of heart disease. By examining and studying the categories and subcategories of mortality of heart disease within New Jersey counties, we can build strategies to make New Jersey a healthier state together.

## Methods

The main outcome of death in adults over 25 from heart disease from 2010 to 2019 was correlated to 28 health factors including comorbidities, behaviors, and socioeconomic factors (see Table 1). Mortality by county from diseases of the heart were pulled from the New Jersey State Health Assessment Data, which in turn was created using CDC data, and was defined as the age-adjusted number of deaths per 100,000 people from coronary artery disease, including heart attacks, arrhythmias, and heart failure from 2010-2019. Mortality figures for each New Jersey county and data from the three counties with the lowest mortality (Hunterdon, Somerset, Bergen) and highest mortality (Cape May, Salem, Cumberland) were used in linear correlation tests in Microsoft Excel with each comorbidity and health factor. This process produced 28 R squared values from the 28 pairs of correlation tests.

**Table 1.** A chart of the comorbidities, behaviors, and socioeconomic factors that were correlated to the outcome, along with descriptions, sources, and the years the data were collected.

| ***Comorbidity*** | **Description** | **Source** | **Year** |
| --- | --- | --- | --- |
| *Obesity* | % of adults reporting BMI >=30 | CDC Diabetes Interactive Atlas | 2015 |
| *Diabetes Mellitus* | Age-adjusted prevalence per 100,000 | CDC Diabetes Interactive Atlas | 2015 |
| *Hypertension* | Age-adjusted prevalence per 100,000 | NJ SHAD | 2010-2019 |
| *Atherosclerosis* | Age-adjusted prevalence per 100,000 | NJ SHAD | 2010-2019 |

| ***Behavior*** | **Description** | **Source** | **Year** |
| --- | --- | --- | --- |
| *Adult smoking* | % of adults who are current smokers | Behavioral Risk Factor Surveillance System | 2016 |
| *Physical inactivity* | % of adults aged 20+ reporting no leisure-time physical activity | CDC Diabetes Interactive Atlas | 2015 |
| *Driving alone to work* | % of workforce driving alone to work | American Community Survey | 2013-2017 |
| *Long commute – driving alone* | Among workers who commute in their cars alone, % commuting >30 minutes | American Community Survey | 2013-2017 |
| *Food Insecurity* | % of population reporting food insecurity | Map the Meal Gap | 2016 |
| *Lack of healthy food* | % of population reporting limited access to healthy food | USDA Food Environment Atlas | 2015 |

| ***Socioeconomic Factor*** | **Description** | **Source** | **Year** |
| --- | --- | --- | --- |
| *Uninsured Adults* | % of adults without health insurance | Small Area Health Insurance Estimates | 2016 |
| *Primary Care Physicians* | Ratio of population to primary care physicians | Area Health Resource File/American Medical Association | 2016 |
| *Children eligible for free lunch* | % of children eligible for free or reduced-price lunch | National Center for Education Statistics | 2016-2017 |
| *Children in poverty* | % of children under age 18 in poverty | Small Area Income and Poverty Estimates | 2017 |
| *Some college* | % of adults 25-44 with some post-secondary education | American Community Survey | 2013-2017 |
| *Single parent household* | % of children that live in a household headed by a single parent | American Community Survey | 2013-2017 |
| *Violent crime* | # of reported violent crime offenses per 100,000 population | Uniform Crime Reporting – FBI | 2014 & 2016 |
| *Fair-to-poor health* | % of adults reporting fair or poor health | Behavioral Risk Factor Surveillance System | 2016 |
| *Poor physical health days* | Average # of physically unhealthy days reported in past 30 days | Behavioral Risk Factor Surveillance System | 2016 |
| *Frequent physical distress* | % of the population reporting frequent physical distress | Behavioral Risk Factor Surveillance System | 2016 |
| *Median household income* | Median household income of the population | Small Area Health Insurance Estimates | 2017 |
| *Rural population* | % of population living in a rural area | Census Population Estimates | 2010 |
| *Life expectancy* | Average life expectancy | National Center for Health Statistics - Mortality Files | 2015-2017 |
| *Age-adjusted mortality* | Overall premature age-adjusted mortality | CDC WONDER mortality data | 2015-2017 |

**Table 2.**
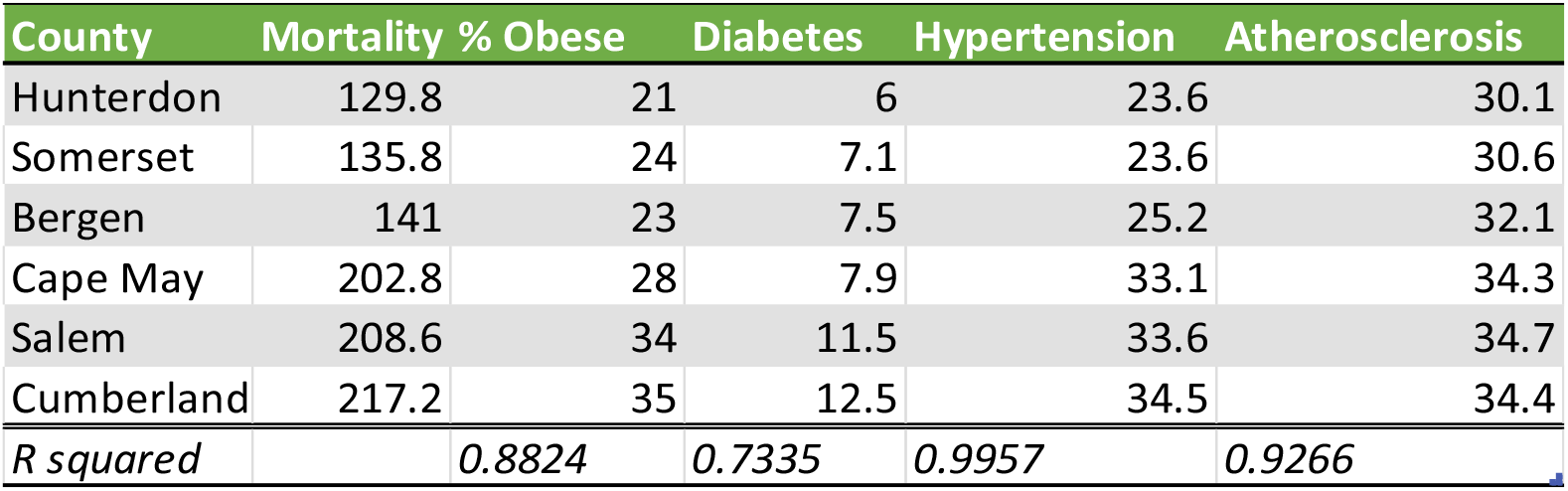
Age-adjusted mortality from diseases of the heart and comorbidities data by county. R squared values are displayed for the linear correlations between mortality and each comorbidity.

**Table 3.** Age-adjusted mortality from diseases of the heart and behaviors data by county. R squared values are displayed for the linear correlations between mortality and each behavior factor.

| County | Mortality | % Smokers | % Phys In | % Drive alone | % Long drive | %Food Ins | % wo healthy food |
| --- | --- | --- | --- | --- | --- | --- | --- |
| Hunterdon | 129.8 | 13 | 18 | 81 | 54 | 6 | 2 |
| Somerset | 135.8 | 11 | 17 | 78 | 46 | 7 | 3 |
| Bergen | 141 | 13 | 21 | 69 | 41 | 8 | 1 |
| Cape May | 202.8 | 15 | 23 | 79 | 24 | 12 | 9 |
| Salem | 208.6 | 17 | 28 | 84 | 37 | 13 | 3 |
| Cumberland | 217.2 | 17 | 30 | 82 | 30 | 13 | 10 |
| *R squared* |  | *0.8492* | *0.8425* | *0.29* | *0.7251* | *0.9823* | *0.5858* |

**Table 4.** Age-adjusted mortality from diseases of the heart and socioeconomic factors data by county. R squared values are displayed for the linear correlations between mortality and each socioeconomic factor.

| County | Mortality | % Uninsured | Pop:PCP | % Free Lunch | % Child Pov | % College |
| --- | --- | --- | --- | --- | --- | --- |
| Hunterdon | 129.8 | 5 | 837 | 9 | 4 | 79 |
| Somerset | 135.8 | 7 | 895 | 21 | 6 | 78 |
| Bergen | 141 | 10 | 839 | 21 | 7 | 77 |
| Cape May | 202.8 | 10 | 1686 | 39 | 16 | 63 |
| Salem | 208.6 | 10 | 2758 | 42 | 22 | 57 |
| Cumberland | 217.2 | 15 | 2330 | 61 | 28 | 40 |
| *R squared* |  | *0.6213* | *0.8599* | *0.8789* | *0.9188* | *0.8481* |
| County | Mortality | % Single Parent | Violence | % Poor Health | Unhealthy Days | % Distress |
| Hunterdon | 129.8 | 15 | 42 | 11 | 2.7 | 8 |
| Somerset | 135.8 | 17 | 64 | 11 | 2.8 | 8 |
| Bergen | 141 | 19 | 84 | 13 | 3.1 | 9 |
| Cape May | 202.8 | 28 | 236 | 15 | 3.6 | 11 |
| Salem | 208.6 | 42 | 266 | 18 | 4.1 | 12 |
| Cumberland | 217.2 | 50 | 516 | 23 | 4.1 | 13 |
| *R squared* |  | *0.8479* | *0.7995* | *0.7855* | *0.9336* | *0.9043* |
| County | Mortality | Income (\$) | % Rural | Life Expectancy | Early Mortalities | |
| Hunterdon | 129.8 | 113083 | 49.6 | 83.1 | 213 |  |
| Somerset | 135.8 | 111838 | 5.8 | 82.5 | 218 |  |
| Bergen | 141 | 93805 | 0.1 | 83.2 | 206 |  |
| Cape May | 202.8 | 64450 | 17.5 | 77.4 | 389 |  |
| Salem | 208.6 | 61322 | 45.3 | 76.7 | 425 |  |
| Cumberland | 217.2 | 51786 | 23 | 75.7 | 452 |  |
| *R squared* |  | *0.9636* | *0.0425* | *0.9851* | *0.9841* |  |

## Results

After 28 R squared values determined between each health factor and the outcome of age-adjusted mortality from diseases of the heart, eight factors were found to correlate extremely well (R^2^ > 0.9) with the outcome. Of the comorbidities, hypertension (R^2^ = 0.9957) and atherosclerosis (R^2^ = 0.9266) prevalence in each county produced the strongest linear relationship with heart disease mortality in a given county. Of the behavioral health factors, the percentage of the population reporting food insecurity (R^2^ = 0.9823) was the strongest correlate with the outcome. Of the socioeconomic factors, overall life expectancy (R^2^ = 0.9851), age-adjusted premature mortality rate (R^2^ = 0.9841), median household income (R^2^ = 0.9636), average reported days spent feeling physically unwell (R^2^ = 0.9336), the percentage of children living in poverty (R^2^ = 0.9188), and the percentage of the population reporting frequent physical distress (R^2^ = 0.9043) produced strong linear correlations with the outcome.

Each of the four comorbidities produced R squared values greater than 0.5 when linear correlation with the outcome was analyzed, compared to five of the six behaviors and thirteen of the fourteen socioeconomic factors. Of the 28 factors analyzed, only two failed to produce R squared values greater than 0.5 when compared to the outcome, which were percentage of the population driving alone to work (R^2^ = 0.29) and the percentage of the population living in a rural area (R^2^ = 0.0425).

## Discussion

Of the factors that strongly correlated with the outcome of mortality from heart disease, some were to be expected simply through a basic understanding of cardiovascular pathogenesis. It is unsurprising that hypertension and atherosclerosis prevalence were strong correlates with heart disease mortality, as those two diseases can contribute to myocardial ischemia and infarctions. For instance, oxidative pathways in the subendothelial space activate pro-inflammatory, immunogenic, and atherogenic processes, result in endothelial dysfunction, plaque growth and destabilization, platelet activation, and thrombosis, ultimately leading to clinical events^13^. Strong linear relationships exist between average reported days spent feeling physically unwell and the outcome and the percentage of the population reporting frequent physical distress and the outcome can be explained by common sense as well, as individuals close to death from a disease of the heart. According to the World Health Organization (WHO) symptoms of cardiovascular disease can include chest pain, shortness of breath, nausea, vomiting, numbness, vision changes, headaches, or loss of consciousness^14^. Additionally, smoking and a poor diet can contribute to pathogenesis of atherosclerosis from a cascade effect caused by oxidized low-density lipoproteins (oxLDL). These lipoproteins act as an antigen that is recognized by macrophages and induces foam cell formation, with ensuing plaque lipid core development, apoptosis, cell death, and cytokine production^15^. As median income, food insecurity and lack of healthy food each correlated well with the outcome, it is likely that poverty and food insecurity lead to difficulty procuring healthy food, which in turn leads to comorbidities such as diabetes and obesity. Diabetic patients account for up to one third of patients in clinical HF trials, with diabetes persisting as an independent predictor of poor outcome^16^. Obesity, which is a significant risk factor for heart disease across sex, age, race/ethnicity, socioeconomic status, and neighborhood environments^17^. Underscoring the role of diet in disease progression, increased intake of fruit and vegetables are associated with a decreased risk of cardiovascular disease^18^. These results place focus on the intersection of diet and income in the progression of heart disease to mortality.

Besides diet, income affects other factors that contribute to mortality such as inability to afford medical appointments and drugs that may stave off death from a cardiac condition. Unfortunately, children burdened with low socioeconomic status bear a disproportionate share of the coronary heart disease burden and coronary heart disease remains the leading cause of mortality in low-income US counties^19^. Child poverty rates are a part of a cluster of developmental factors surveyed within the group of socioeconomic factors. In addition to growing up in a single parent household, receiving free or reduced-price lunches at schools, and not attending college, these data suggest that the road to death from heart disease begins well before adulthood, and serve to underscore the importance of public health interventions in childhood and economic opportunities for working families. Populations with upward mobility (versus consistent low socioeconomic status) from childhood to adulthood were associated with a greater prevalence, but lower incidence of hypertension^20^.

The results also provide room to discuss the importance of primary care in the prevention of heart disease mortality. The data suggested that the more primary care physicians that practice in a county, the less that county experiences death from diseases of the heart. Primary care physicians can act as a first line of defense in mortality from heart disease because they can prescribe certain drugs to begin to protect the patient’s heart health. A possible solution to expand access to primary care in underserved communities and alleviate overwhelmed physicians is practice co-management with nurse practitioners^21^. This can ultimately buy time for the patient to see a specialist if needed. Following in this vein, higher percentages of uninsured people in the population correlated with increased mortality as well, possibly from the uninsured avoiding preventive interventions or having trouble affording beneficial medications. This data affirms the continued need for affordable and effective healthcare in at-risk communities, such as the NIH’s National Heart Lung and Blood Institute’s guidelines that could potentially prevent about 56,000 cardiovascular events and 13,000 deaths annually, while saving costs^22^.

Nearly every factor produced a linear relationship with the outcome with an R squared value greater than 0.5, the threshold at which the presence of a correlation is considered, save for two. The percentage of the population living in a rural area did not correlate with the outcome. This was somewhat surprising, as the three counties with the highest mortality from diseases of the heart had, on average, a greater percentage of their population living in rural areas. And while the percentage of the population driving alone to work did not correlate with the outcome, interestingly, the percentage of lone drivers reporting a long commute did. This may be a tie-in to the percentage of the population reporting a sedentary lifestyle, as long commutes require sitting for extended periods of time. Although this provides positive data, more additional investigation should be done into the availability of opportunities for exercise during everyday life, especially those in disadvantaged communities. Disadvantaged communities throughout New Jersey should be further studied for walkability and it should be determined if walking paths or sidewalks are available in such communities. The role of public safety cannot be ignored either, as living in an area with higher amounts of violent crime correlated with increased mortality from heart disease as well, possibly by creating an unsafe outdoor environment and discouraging jogging and other outdoor exercise.

Limitations including the potential presence of confounding variables must be identified and minimized to the furthest extent possible. Admittedly, due to the overarching impact of socioeconomic status across all communities and risk factors, this will be difficult, if not impossible, to do. For example, the odds of adults being overweight were 30% higher in individuals living in low socioeconomic status neighborhoods, compared with that of individuals living in high socioeconomic status neighborhoods^23^. This blurs the line between comorbidities and the role of social factors in mortality. As overall average life expectancy and age-adjusted premature mortality correlate with the outcome as well, further work will be done to determine which factors, if any, correlate specifically to mortality from heart disease, rather than deaths from cancer, for instance. Additionally, some of the data used in this study were collected at just one point over the ten-year period of study. The general lack of routinely reported information on social and economic differences in health may or may not have impacted the validity of this study, but certainly has public health implications^24^.

Other future directions include using these factors to create a model that should be able to accurately predict the mortality from diseases of the heart in New Jersey. This model would help us to identify the top five predictive factors for mortality from diseases of the heart. The goal would be for the model to predict mortality after the data for the health factors from a county was inputted. If the model was able to accurately complete this task, then it could be assumed that it could be used to predict future mortality based on anticipated changes in health factors. It is understood among the socioeconomic factors, mainly race/ethnicity and marital status affect the risk for readmission in elderly people with heart failure or acute myocardial infarction^25^. It will be interesting to see what future models may determine as leading factors. This information would be of immense use to the public health community, as they would be able to create more targeted interventions in the future.

## Conclusion

This paper examines six counties in New Jersey that were divided into the least (Hunterdon, Somerset, Bergen) and greatest (Cape May, Salem, and Cumberland) mortality rates from heart disease. The overall results suggest that mortality from heart disease is not caused by a single factor, but by multiple interconnected factors. The comorbidity data that correlated hypertension and atherosclerosis produced the strongest linear relationship with mortality of heart disease mortality per county. Of the behavior patterns data, food insecurity produced the strongest correlation with the outcome. However, the socioeconomic status category was more complicated in which multiple subcategories produced strong correlations that included overall life expectancy, age-adjusted premature mortality rate, median household income, average reported days spend feeling physically unwell, the percentage of children living in poverty, and the percentage of the population reporting frequent physical distress. Through education and advocacy work, major strides can be taken to tackle the mortality of heart disease crises and begin implementing public health strategies to make all of New Jersey a healthier state for all communities.

## Data Availability

All data produced in the present work are contained in the manuscript

## Acknowledgments

None

## Notes

### Competing Interest Statement

The authors have declared no competing interest.

### Funding Statement

This study did not receive any funding

